# The contribution of Apoliproprotein E genetic variation to dementia risk in British South Asians

**DOI:** 10.1101/2025.09.04.25334903

**Authors:** Benjamin M Jacobs, Avinash Chandra, Arnab Mandal, Isabelle Foote, Faiza Durrani, Sheena Waters, Yue Liu, Petroula Proitsi, Genes & Health Research Team, Cara L. Croft, Dylan M Williams, Moneeza Siddiqui, Sarah Finer, Stuart Rison, David A van Heel, Charles R Marshall

## Abstract

**INTRODUCTION:** Understanding the genetic basis of dementia in diverse populations is essential to ensure that efforts to predict, prevent, and treat dementia are equitable. The strongest genetic risk factor for dementia - *APOE* genotype - has not been assessed in population-scale cohorts of South Asian ancestry.

**METHODS:** We analysed data from 51,104 volunteers in the Genes & Health study - a cohort study of British South Asians - who have undergone genotyping and consented for linkage to healthcare records. All-cause dementia was defined using electronic healthcare records. *APOE* genotypes were defined using phased, imputed genotype data. Cox proportional hazards models were used to assess the relationship between *APOE* genotype and dementia. Population attributable fractions were calculated for each *APOE* genotype.

**RESULTS:** *APOE _ε_*4 was associated with all-cause dementia in a dose-dependent fashion (Case N = 614, Control N = 50,490; *APOE* _ε_4/_ε_4: Hazard Ratio 2.7, *P* < 0.0001; *APOE* _ε_4/_ε_3: Hazard Ratio 1.5, *P* < 0.001). The overall proportion of dementia cases attributable to this allele was 14.2% (95% CI -2.8% - 25.0%). *APOE* _ε_4 was also associated with elevated triglycerides and Low Density Lipoprotein (LDL) cholesterol.

**DISCUSSION:** *APOE* _ε_4 - the major genetic risk factor for sporadic dementia in European-ancestry populations - has a similar impact on dementia risk in British South Asians.

## Research in context

### Systematic review

We reviewed the literature using traditional sources (e.g. Pubmed) for articles containing terms ‘APOE’, ‘dementia’ or ‘Alzheimer/s’, and ‘ancestry’ or ‘South Asian’ in the title. Although *APOE* has been extensively studied in European-ancestry cohorts, and more recently ancestrally diverse cohorts such as AllOfUs and Million Veterans Programme, few studies have explored the impact of *APOE* on dementia risk in biobank-scale cohorts of South Asian ancestry.

### Interpretation

We show that the common *APOE* alleles _E_4 and _E_3 are associated with increased dementia risk in a British South Asian cohort. At a population-level, these alleles are likely to explain a comparable overall proportion of dementia cases.

### Future directions

Prediction and prevention studies aiming to use *APOE* to identify people at high risk of dementia should be inclusive of all ancestries. Genetic analysis of dementia risk in diverse populations, including South Asian cohorts, is essential for ensuring that the downstream benefits - for prediction, prevention, and drug discovery - are shared equitably.

## Introduction

Genome-wide association studies (GWAS) of sporadic Alzheimer’s disease (AD) have demonstrated that AD has a strong genetic component driven by common variation in at least 75 risk loci^1,2^. Variation at the *APOE* locus (encoding apolipoprotein E; APOE) alone is estimated to account for over one third of cases^3^, however this is based almost entirely on evidence from those of European ancestry.

Common coding variants in *APOE* define three common APOE protein isoforms - _E_2, _E_3, and _E_4 - of which _E_4 is associated with increased risk of AD in a dose-dependent fashion and _E_2 with decreased risk with respect to the most common allele, _E_3^4,5^. The association between haplotypes at the *APOE* locus and AD has been consistently demonstrated across cohorts^1,6,7^, including populations of European, recently admixed (African American), Hispanic, and East Asian ancestry. The association between *APOE* genetic variation and dementia risk has not been evaluated at scale in a population of South Asian ancestry, although small studies in Pakistani and Indian populations have suggested an effect similar to that seen in European populations^8–12^. British South Asians are at higher dementia risk and tend to be diagnosed with dementia at a younger age ^13,14^, experience mortality sooner after a dementia diagnosis^13^, and have higher prevalence of dementia risk factors including deprivation, dyslipidaemia and type 2 diabetes mellitus^15,16^. Understanding the genetic basis of dementia in this population is essential for prediction and prevention efforts, especially given a surge in forecasted dementia cases in coming decades^17^. To date, few studies have examined the prevalence of *APOE* variants and the magnitude of their associations with dementia in a population-scale cohort of South Asian ancestry.

We therefore aimed to consider whether the major European-ancestry genetic risk factor for sporadic dementia - *APOE* genotype - was associated with dementia and other health-related phenotypic outcomes in a cohort of ∼70,000 British South Asian participants in the Genes & Health study. We hypothesised that both allele frequencies and effects on dementia risk might vary relative to European ancestry populations.

## Methods

### Cohort

Genes & Health (G&H) is a longitudinal cohort study of self-identified British Bangladeshi and British Pakistani individuals with genetic data (array genotyping and exome sequencing) and linked electronic healthcare records for over 70,000 healthy volunteers, with recruitment expected to expand to over 100,000 individuals^16^.

### Genetic data & Definition of *APOE* haplotypes

G&H volunteers were genotyped from saliva using the Illumina Global Screening Array version 3.0. Genotype quality control procedures have been previously described^18–20^. *APOE* haplotypes were determined from phased genotype data imputed to the Topmed-r3 reference^21^. Genotypes at two coding variants - rs429358 and rs7412 - define the common *APOE* haplotypes^22^ as follows: rs429358T - rs7412T (_E_2), rs429358T - rs7412C (_E_3), rs429358C - rs7412C (_E_4). We used data from the October 2023 data freeze, comprising imputed genetic data for 51,166 volunteers and excluded 62 volunteers of ambiguous genetic ancestry, giving a final sample size of 51,104.

### Phenotype definitions

All-cause dementia cases were identified using a custom codelist applied to multiple sources of linked electronic healthcare records, including NHS Digital Hospital Episode Statistics, primary care data, and Barts Health and Bradford Teaching Hospitals secondary care records. All-cause dementia was used as the primary outcome as this both maximises the sample size (and thus statistical power) and because more precise, clinically-subtyped diagnoses of dementia phenotypes are generally unreliable in electronic healthcare records^23^. In secondary sensitivity analyses we used Alzheimer’s Dementia (ICD code G30 or F00), unspecified dementia (ICD code F03) and vascular dementia (F01) as the outcome measure. Phenotypes were curated as part of a bespoke Python pipeline. All code and codelists used for phenotype generation are available at https://github.com/genes-and-health/BI_PY.

### Statistical analysis

#### Demographics

Demographic data are presented with medians and interquartile ranges for continuous variables and as numbers and percentages for categorical variables.

#### Allele frequency comparison

We tested for statistical enrichment/depletion of the *APOE* _E_4 allele in the Genes & Health cohort using a two-tailed binomial test, comparing the proportion of _E_4 carriers and non-carriers with the proportion of carriers observed in the subset of ∼340,000 White British UK Biobank volunteers^24^.

#### Association of *APOE* haplotypes with dementia

Survival analysis was conducted using Cox proportional hazards models. As individuals were recruited from the age of 16 or over, the ‘time-to-event’ was defined as the time from the age of 16 to the age of dementia diagnosis. For controls, the censoring time was defined as December 2024 when the last phenotype data extract was performed. Models were adjusted for age, gender, the first ten genetic principal components, and binary genetic ancestry (inferred Pakistani or Bangladeshi ancestry). For models, the _E_3/_E_3 allele was used as the reference as this is the most common allele. Each pair of alleles was considered separately (i.e. a ‘genotypic’ model). Hazard ratios are expressed with 95% confidence intervals and *P* values for the null hypothesis that the ratio = 1. Models were inspected for the proportional hazards assumption using Schoenfeld residuals and for linearity. As sensitivity analyses, we repeated the analysis using fewer covariates (age and gender alone), using logistic regression models instead of Cox models (adjusted for identical covariates), we used gender-stratified models, we repeated the analysis using coded ‘Alzheimer’s dementia’ rather than all-cause dementia, and we restricted the analysis to those recruited over the age of 60 and excluded cases diagnosed prior to 60 (note for these models age 60, rather than age 16, was used as time zero for survival analyses).

#### Population attributable fraction (PAF)

We estimated the PAF for all-cause dementia by using the stratified exposure methods described in Hanley (2001)^26^ and used in Williams *et al.* (2025)^3^. To do so we first used a logistic regression model of the form dementia ∼ *APOE* genotype + covariates, using the low-risk, rare _ε_2/_ε_2 genotype as reference, to derive as estimate of the Odds Ratio for dementia given each combination of *APOE* alleles. This means individuals with intermediate risk genotypes (carriers of _ε_3) are not subsumed into the reference group, which would be expected to bias downwards proportions attributable to risk alleles. We calculated the ‘case fraction’ i.e. the proportion of all cases accounted for by each genotype (i.e. N cases / total cases), and estimated the genotype-specific PAF as (OR - 1) / (OR x case fraction). The overall PAF attributable to either _ε_3 or _ε_4 was estimated by summing the genotype-specific PAFs overall all genotypes containing _ε_3 or _ε_4 (_ε_3/_ε_3, _ε_3/_ε_4, _ε_3/_ε_2, _ε_4/_ε_2, _ε_4/_ε_4). To estimate the PAF attributable to _ε_3 or _ε_4 alone, we decomposed the relative contributions of _ε_3 and _ε_4 to the effect estimate for the _ε_3/_ε_4 genotype using the approach described in Williams *et al.* (2025)^3^; specifically, the ratio of the Odds Ratios for _ε_4/_ε_2 and _ε_3/_ε_2 was used to derive a weight (ratio / ratio + 1 for _ε_4; 1 - this weight for _ε_3) quantifying the proportion of the PAF for _ε_4/_ε_3 attributable to each allele individually.

Confidence intervals were calculated by calculating the genotype-specific PAF for each stratum using the lower or upper bound of the effect estimate (OR +/- 1.96 x standard error), and summing these PAFs together. The confidence estimates for the decomposed effect of _ε_3/_ε_4 were derived using the upper and lower bounds of the ratio.

#### Phenome-wide association study

We analysed the association of *APOE* alleles relative to _ε_3/_ε_3 with all available binary (disease) traits with at least 500 cases and all quantitative traits with N > 1000. We used logistic (for binary traits) and linear (for quantitative) traits adjusted for the same covariates as in the primary Cox models (age, gender, first ten PCs, and ancestry). In the text we report associations surpassing a study-wide Bonferroni threshold correction to maintain an alpha of 5% (i.e. *P* = 0.0002 = 0.05 / 236 tests).

## Results

### *APOE* **ε**4 prevalence in British South Asians

We analysed genetic and healthcare data from 51,104 British South Asian participants in the Genes & Health study (**methods**). Demographic characteristics of identified dementia cases and controls are shown in the table (**table 1**). Both variants used to define *APOE* alleles were common and well-imputed in G&H: rs429358 (chr19:44908684:T:C, Minor allele frequency [MAF] = 0.10, Imputation quality score [INFO] = 0.96); rs7412 (chr19:44908822:C:T, MAF = 0.05, INFO = 0.92). For both of these *APOE* variants, the allele frequency was consistent with estimates from reference populations of South Asian (SAS) ancestry but lower than in Non-Finnish European (NFE) reference samples (rs429358: MAFG&H = 10%, MAFgnomAD-SAS = 10.1%^27^, MAFgnomAD-NFE = 15.1%; rs7412 : MAFG&H = 5%, MAFgnomAD = 4.2%, MAFgnomAD-NFE = 7.8%). The overall allele frequencies for *APOE* haplotypes in G&H were 10.4% for *APOE* _ε_4, 85.1% for *APOE* _ε_3, and 4.5% for *APOE* _ε_2. The allele frequency of *APOE* _ε_4 was 11.3% in the participants of inferred Bangladeshi ancestry and 9.1% in participants of inferred Pakistani ancestry.

**Table 1:** demographic characteristics.

|  | Dementia | Controls |
| --- | --- | --- |
| **N** | 614 | 50,490 |
| **Age at recruitment (median, [IQR])** | 71.3 [15.9] | 39.0 [17.5] |
| **Gender (n, %)** |  |  |
| Female | 254 [41.4%] | 28,018 [55.5%] |
| Male | 260 [58.6%] | 22,472 [44.5%] |
| **APOE genotype** |  |  |
| $\epsilon 2/\epsilon 2$ | <10 [<1.7%] | 151 [0.3%] |
| $\epsilon 2/\epsilon 3$ | 42 [6.8%] | 3,794 [7.5%] |
| $\epsilon 2/\epsilon 4$ | <10 [<1.7%] | 475 [0.9%] |
| $\epsilon 3/\epsilon 3$ | 412 [67.1%] | 36,710 [72.7%] |
| $\epsilon 3/\epsilon 4$ | 131 [21.3%] | 8,776 [17.4%] |
| $\epsilon 4/\epsilon 4$ | 20 [3.3%] | 584 [1.2%] |
| **APOE allele frequency** |  |  |
| $\epsilon 2$ | 4.2% | 4.5% |
| $\epsilon 3$ | 81.2% | 85.2% |
| $\epsilon 4$ | 14.6% | 10.3% |
| **Inferred genetic ancestry** |  |  |
| Bangladeshi | 335 [54.6%] | 29,614 [58.7%] |
| Pakistani | 279 [45.4%] | 20,876 [41.3%] |
| **Age at diagnostic code (median, IQR)** | 73.3 [16.0] |  |

Dementia cases (n = 614, of which 451 [73.5%] were incident) were recruited at an older age (median 71.3 vs 39.0), were more likely to be male (58.6% vs 44.5%), and were more likely to carry at least one *APOE* _ε_4 allele (25.9% vs 19.5%) than controls without dementia (n = 50,490). Overall, 19.6% of the population carried at least one *APOE* _ε_4 allele (18.4% heterozygous, 1.2% homozygous), and 97.5% carried at least one *APOE* _ε_3 allele (24.9% heterozygous, 72.6% homozygous). The proportion of British South Asians who carry at least one *APOE* _ε_4 allele is therefore somewhat lower than White British people (19.6% vs 28.8% in UK Biobank^24^, two-tailed Binomial test *P* < 2x10^-16^), but consistent with prior estimates from cohorts of South Asian ancestry (e.g. 19.4% in an Indian population^8^ and 18.4% among the few self-identified British South Asians in UK Biobank^28^).

### Association of *APOE* alleles with dementia in Genes & Health

Carriage of the *APOE* _ε_4 allele was associated with dementia diagnosis in a dose-dependent fashion (**figure 2**; *APOE* _ε_4/_ε_4: Hazard Ratio [HR] 2.7, 95% CI 1.7 - 4.2, *P* < 0.0001; *APOE* _ε_4/_ε_3: Hazard Ratio [HR] 1.5, 95% CI 1.2 - 1.8, *P* < 0.001; Cox multivariable regression models adjusted for age at recruitment, gender, PCs 1 - 10, and genetic ancestry; all models used *APOE* _ε_3/_ε_3 as reference).

**Figure 1:**
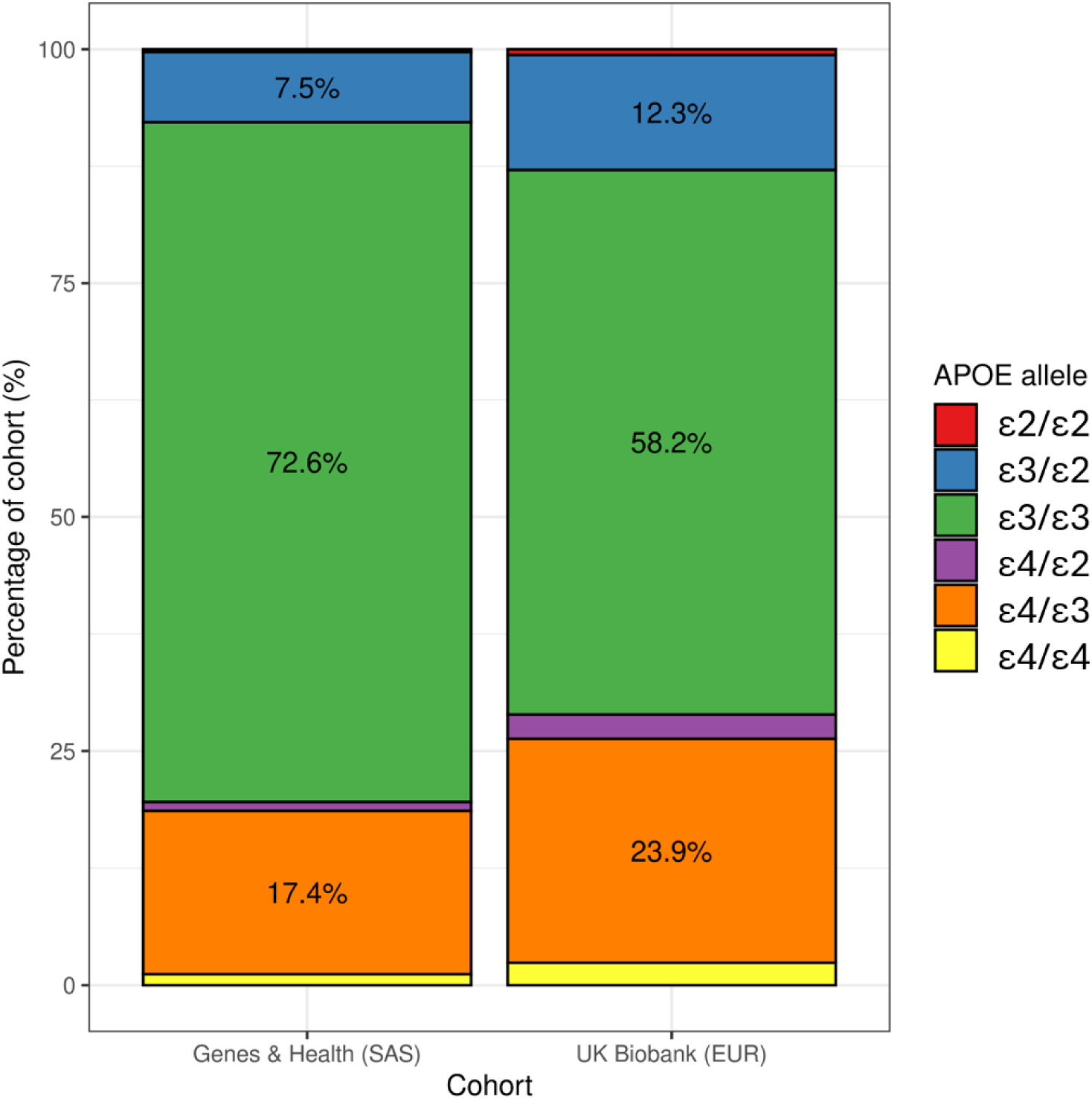
*APOE* genotype prevalence in British South Asians. Stacked barplots show the distribution of *APOE* genotypes (proportions) for the Genes & Health cohort (left), comprising British Bangladeshi and British Pakistani volunteers, compared with UK Biobank participants of European ancestry (right). The components of the barplot are coloured according to inferred *APOE* genotype. Percentage labels are shown for percentages > 5%. SAS - South Asian ancestry. EUR - European ancestry.

**Figure 2:**
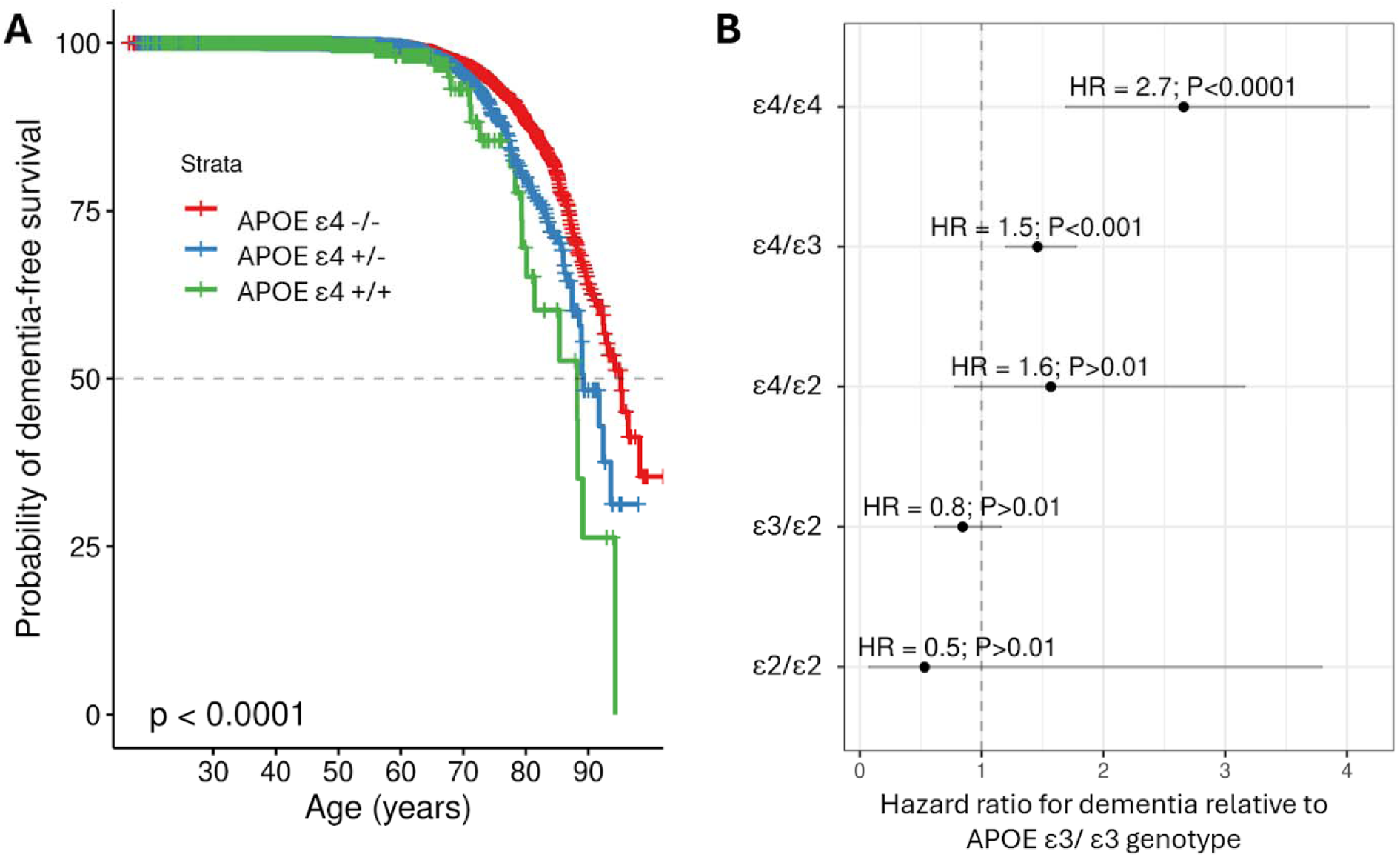
Association between *APOE* alleles and all-cause dementia in ∼50,000 British South Asians. B - survival curves indicating the probability of dementia-free survival in carriers of 0, 1, or 2 *APOE* e4 alleles. C - forest plot indicating the Hazard Ratios and 95% confidence intervals for the association of each combination of *APOE* alleles with all-cause dementia, with the _ε_3/_ε_3 genotype as the reference.

We observed weaker (i.e. *P* > 0.01) evidence for a risk-increasing association of *APOE* _ε_4/_ε_2 (HR 1.6, 95% CI 0.8 - 3.2, *P* = 0.2) and protective association of the rarer _ε_2 genotypes *APOE* (_ε_2/_ε_2: HR 0.5, 95% 0.1 - 3.8, *P* =0.53; _ε_2/_ε_3: HR 0.8, 95% 0.6 - 1.2, *P* =0.3). Each copy of the *APOE* _ε_4 allele was associated with a 55% increase in the hazard of all-cause dementia (HR 1.56, 95% CI 1.34 - 1.82, *P* = 1.1x10^-8^, additive genetic model). In a dominant model, carriage of the *APOE* _ε_4 allele was associated with a similar degree of risk increase (HR 1.58, 95% CI 1.32 - 1.90, *P* = 7.3x10^-7^). The magnitude of this association strengthened when considering Alzheimer’s disease rather than all-cause dementia as the outcome (*APOE* _ε_4/_ε_4: Hazard Ratio 7.4, 95% CI 3.1 - 17.6, *P* < 0.0001; *APOE* _ε_4/_ε_3: Hazard Ratio 2.2, 95% CI 1.3 - 3.7, *P* = 0.002) although these estimates were imprecise owing to the smaller number of cases (NAD = 82).

The association between *APOE* status and all-cause dementia was similar in men and women: in gender-stratified models, *APOE* _ε_4 homozygosity was associated with a substantially elevated risk in both males (HR 2.73, 95% CI 1.55 - 4.80) and females (HR 2.95, 95% CI 1.38 - 6.33). We observed a similar magnitude of association using logistic regression models (_ε_4/_ε_4: OR 3.03, 95% CI 1.8 - 5.2, *P* = 7.3x10^-7^; _ε_4/_ε_3: OR 1.47, 95% CI 1.2 - 1.8, *P* = 0.0004). As expected, considering the rare _ε_2/_ε_2 genotype as the reference genotype enhanced the effect sizes of genotypes containing _ε_3 and _ε_4 alleles, while reducing the precision of the estimates (e.g. _ε_4/_ε_4: OR 7.46, 95% CI 0.86 - 65.0, *P* = 0.07) due to the rarity of this genotype.

To account for bias due to the younger age of the control cohort, we performed a sensitivity analysis restricting to participants recruited over the age of 60 and dementia cases diagnosed >60, yielding a subset of 484 dementia cases (median age at recruitment 74.8, IQR 11.8; median age at diagnostic code report 76.6, IQR 12.7; 39.5% female) and 5,015 controls (median age at recruitment 66.0, IQR 9.0; 45.8% female). The association results were similar to the primary analysis (*APOE* _ε_4/_ε_4: Hazard Ratio [HR] 2.8, 95% CI 1.7 - 4.6, *P* < 0.0001; *APOE* _ε_4/_ε_3: HR 1.6, 95% CI 1.3 - 2.0, *P* = <0.0001; *APOE* _ε_4/_ε_2: HR 2.1, 95% CI 1.04 - 4.27, *P* = 0.04; *APOE* _ε_3/_ε_2: HR 0.9, 95% CI 0.6 - 1.2, *P* = 0.42; *APOE* _ε_2/_ε_2: HR 0.6, 95% CI 0.1 - 4.6, *P* = 0.66).

### Population attributable fraction

To quantify the population-level significance of *APOE* variants for dementia in this population we estimated the population attributable fraction (PAF) for all-cause dementia (**figure 3**). We estimated the PAF for all-cause dementia contingent on the presence of any risk-increasing *APOE* allele (i.e. _ε_3 or _ε_4) as 62.6%, however due to the scarcity of _ε_2/_ε_2 genotypes th confidence intervals were broad (95% CI -206% - 95.4%). We estimated that approximately 14.2% of all-cause dementia cases could be attributed to the presence of *APOE* _ε_4 (95% CI - 2.8% - 25.0%), and 48.4% (95% CI -203% - 70.3%) to *APOE* _ε_3.

**Figure 3:**
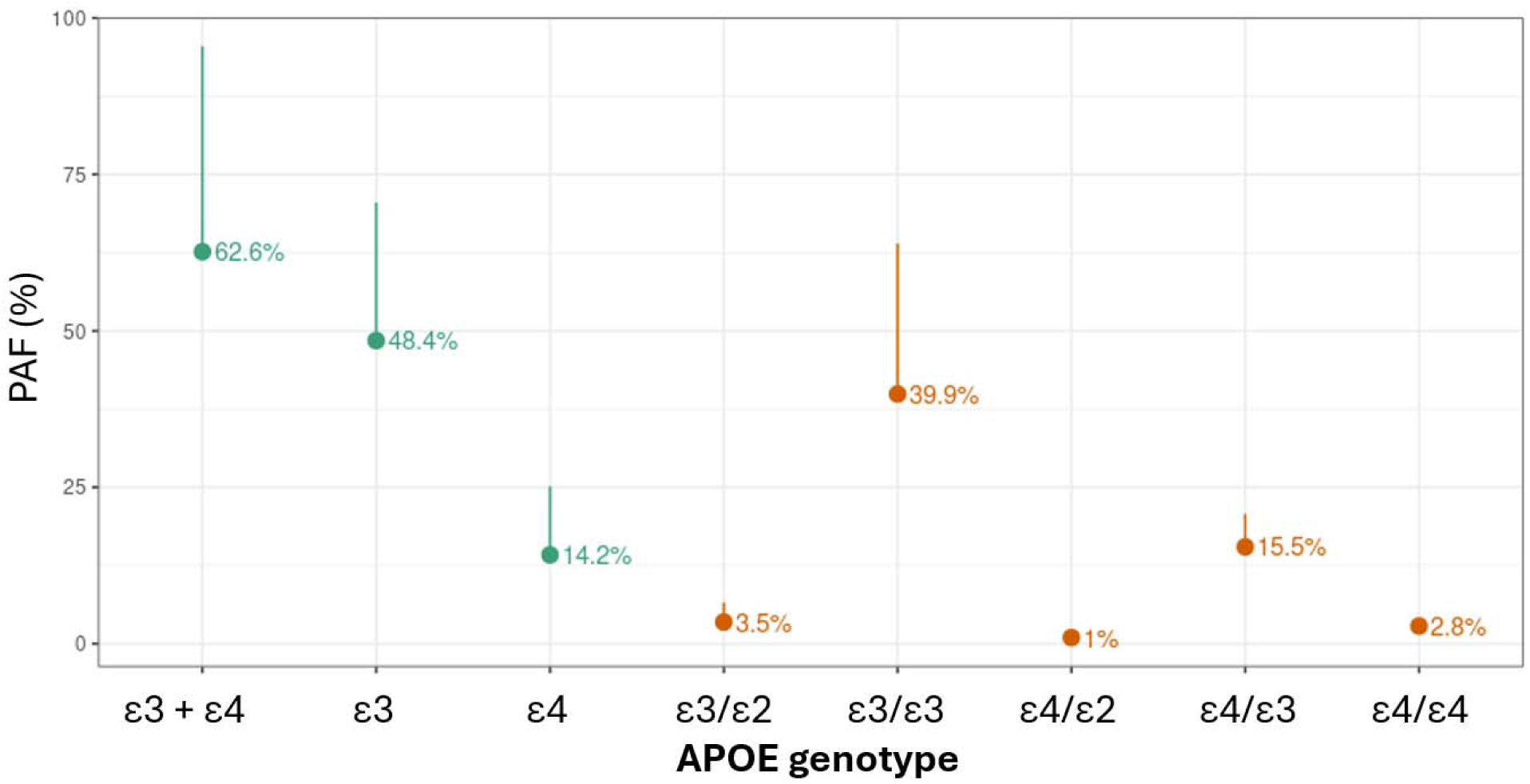
Population attributable fractions (PAF) for *APOE* alleles. Points in orange indicate the estimated PAF for each combination of *APOE* genotypes, relative to the low risk ε2/ε2 reference category. Points in dark green indicate the ‘summed’ PAFs, summarising either the contribution of ε4 alleles only, ε3 alleles only, or the joint PAF attributable to both ε4 and ε3. The upper 95% confidence interval is shown. The lower 95% confidence interval is not shown for clarity, as some estimates were imprecise (see text) due to the rarity of the reference genotype and hence large standard errors attached to the estimates.

### Phenome-wide associations of *APOE*

Dementia-associated *APOE* haplotypes (*APOE* _ε_4 and _ε_3) were associated with higher triglycerides, Low Density Lipoprotein (LDL) cholesterol, and total cholesterol, and with lower CRP (Bonferroni-adjusted *P* value < 0.05, total number of traits tested = 236). Other than for these traits and dementia, no other associations persisted at phenome-wide significance (**figure 4**).

**Figure 4:**
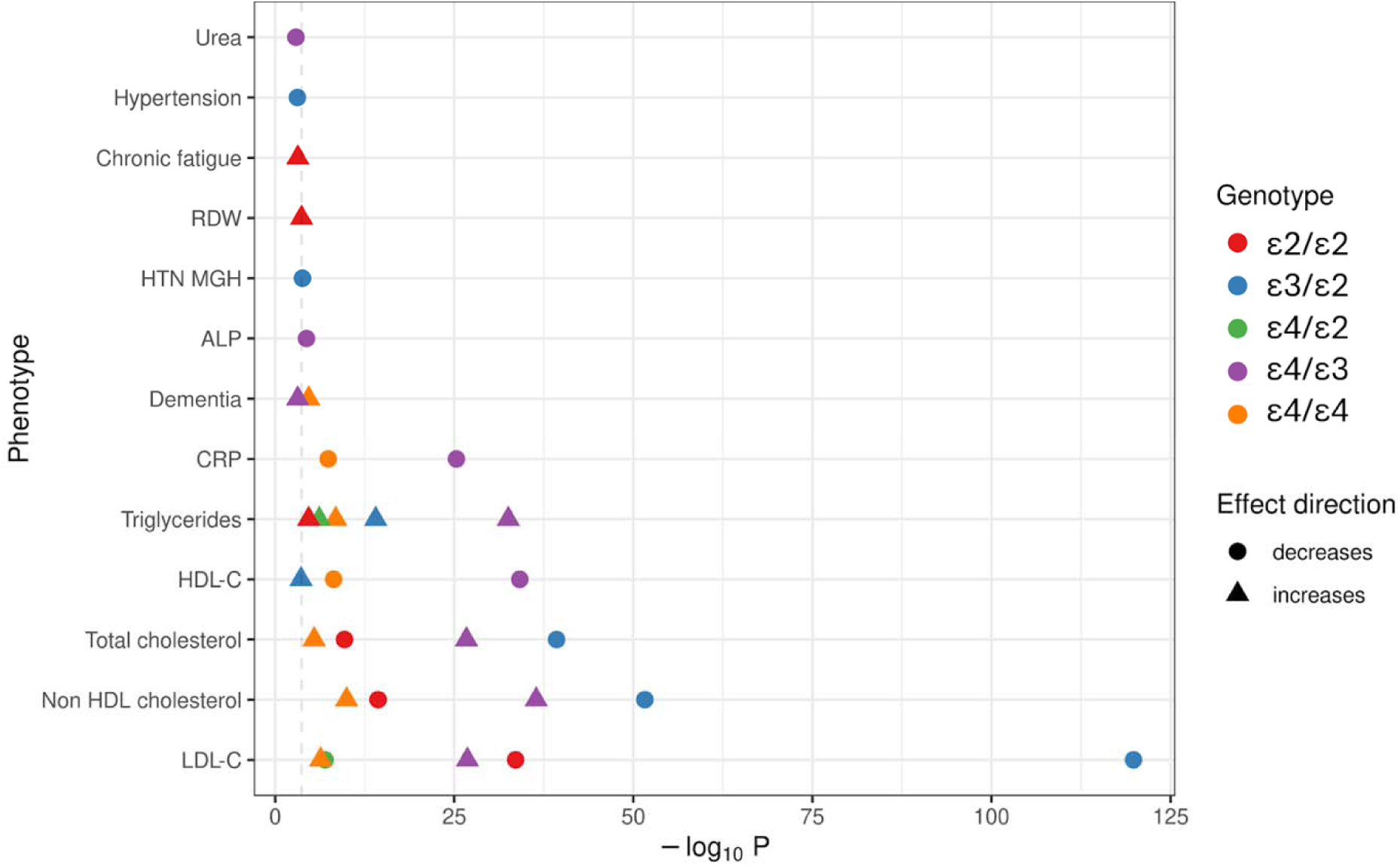
Phenome-wide associations of *APOE*. The plot shows the association (from linear or logistic regressio models) between each *APOE* genotype and each of the tested quantitative and binary traits in Genes & Health (n = 236 traits). Points are shown for genotype-trait associations surpassing a False Discovery Rate of 5%. The dashed line indicates the Bonferroni threshold of 5%. The shape of the point indicates the effect direction, and the x axis indicates the strength (i.e. -log10[P]). The colour indicates the genotype. All effects are orientated to the reference _ε_3/_ε_3 allele. RDW: Red Cell Distribution Width. HTN MGH: Hypertension, alternative codelist definition. ALP: Alkaline Phosphatase. CRP: C-Reactive Protein. HDL-C: High Density Lipoprotein Cholesterol. LDL-C: Low Density Lipoprotein Cholesterol.

## Discussion

Here we demonstrate that the major genetic risk factor for sporadic Alzheimer’s Dementia - *APOE* variation - is associated with all-cause dementia in a biobank-scale cohort of South Asian ancestry, Genes & Health. Specifically, we show that the dementia-associated *APOE* _ε_4 allele i associated with all-cause dementia in a dose-dependent manner, with homozygotes almost 3 times as likely to develop dementia as those with the most common _ε_3/_ε_3 genotype. The population attributable fraction for dementia contingent on the presence of either *APOE* _ε_3 or _ε_4 - the proportion of cases which could theoretically be prevented by neutralising the impact of these variants - was estimated at 62.6%, analogous to estimates from predominantly White British UK Biobank participants and other predominantly European-ancestry cohorts^3^.

In line with the slightly lower frequency of the _ε_4 allele in this population than in Europeans, the estimated PAF for _ε_4 alone is slightly lower than European-ancestry estimates (14.2% in Genes & Health vs 29.3% in UK Biobank - unpublished data, Dylan WIlliams), although the broad confidence intervals mandate caution in interpreting this difference. Furthermore, we show that the association between *APOE* variants and dementia does not differ substantially by gender, and we replicate the association of *APOE* _ε_3 and _ε_4 with elevated triglycerides and LDL cholesterol as expected from previous work^24^.

The major limitations of this work are the relatively young age of the cohort and hence risk of incomplete ascertainment (many participants who will develop dementia in the future have not yet developed it) and the phenotyping of dementia cases, which relies on electronic healthcare records and may therefore mix various pathological subtypes of dementia, including misdiagnoses, and does not allow for a detailed examination of dementia phenotypes. Our use of imputed genetic data rather than sequencing is a limitation, but the very high imputation quality and the fact that both markers used are common is reassuring. Further work with sequencing data will be required to evaluate *APOE* haplotypes beyond the three two-SNP haplotypes examined in this paper, as there may be population-enriched variants seen only in this population which modulate dementia risk in addition to the SNPs encoding the major APOE isoforms^29,30^.

Our findings confirm *APOE* _ε_4 and _ε_3 as genetic risk factors for dementia in people of South Asian ancestry and suggest that at the population-level, these genetic factors alone account for a sizeable proportion of dementia cases. However, _ε_4 prevalence, effect size and PAF all showed a trend towards being lower in this British South Asian population than in those of European ancestry, suggesting that *APOE* variation is does not fully account for the excess dementia risk that we have previously described in this population^14^.

## Data availability statement

All code used to produce these results is available at https://benjacobs123456.github.io/apoe_gh/.

## Funding information

BMJ is supported by a Guarantors of Brain post-doctoral fellowship award and an NIHR Clinical Lectureship. AC is funded by the National Institute for Health and Care Research (NIHR; ref: NIHR203373). This work was supported by an Alzheimer’s Research UK (ARUK) London Network Centre Pump Prime funding award. SW is supported by the Global Parkinson’s Genetics Program (GP2). GP2 is funded by the Aligning Science Across Parkinson’s (ASAP) initiative and implemented by The Michael J. Fox Foundation for Parkinson’s Research (https://gp2.org).

## Data Availability

All code used to produce these results is available at https://benjacobs123456.github.io/apoe_gh/.

## Acknowledgements

Genes & Health is/has recently been core-funded by Wellcome (WT102627, WT210561), the Medical Research Council (UK) (M009017, MR/X009777/1, MR/X009920/1), Higher Education Funding Council for England Catalyst, Barts Charity (845/1796), Health Data Research UK (for London substantive site), and research delivery support from the NHS National Institute for Health Research Clinical Research Network (North Thames). We acknowledge the support of the National Institute for Health and Care Research Barts Biomedical Research Centre (NIHR203330); a delivery partnership of Barts Health NHS Trust, Queen Mary University of London, St George’s University Hospitals NHS Foundation Trust and St George’s University of London.

This research was conducted as part of the ‘Brain Consortium’ approved project (ID S00015).

Genes & Health is/has recently been funded by Alnylam Pharmaceuticals, Genomics PLC; and a Life Sciences Industry Consortium of AstraZeneca PLC, Bristol-Myers Squibb Company, GlaxoSmithKline Research and Development Limited, Maze Therapeutics Inc, Merck Sharp & Dohme LLC, Novo Nordisk A/S, Pfizer Inc, Takeda Development Centre Americas Inc.

We thank Social Action for Health, Centre of The Cell, members of our Community Advisory Group, and staff who have recruited and collected data from volunteers. We thank the NIHR National Biosample Centre (UK Biocentre), the Social Genetic & Developmental Psychiatry Centre (King’s College London), Wellcome Sanger Institute, and Broad Institute for sample processing, genotyping, sequencing and variant annotation. This work uses data provided by patients and collected by the NHS as part of their care and support. This research utilised Queen Mary University of London’s Apocrita HPC facility, supported by QMUL Research-IT, http://doi.org/10.5281/zenodo.438045

We thank: Barts Health NHS Trust, NHS Clinical Commissioning Groups (City and Hackney, Waltham Forest, Tower Hamlets, Newham, Redbridge, Havering, Barking and Dagenham), East London NHS Foundation Trust, Bradford Teaching Hospitals NHS Foundation Trust, Public Health England (especially David Wyllie), Discovery Data Service/Endeavour Health Charitable Trust (especially David Stables), Voror Health Technologies Ltd (especially Sophie Don), NHS England (for what was NHS Digital) - for GDPR-compliant data sharing backed by individual written informed consent.

Most of all we thank all of the volunteers participating in Genes & Health.

A favourable ethical opinion for the main Genes & Health research study was granted by NRES Committee London - South East (reference 14/LO/1240) on 16 Sept 2014. Queen Mary University of London is the Sponsor, and Data Controller.

## Competing interests

The authors declare no relevant competing interests.

## Notes

### Competing Interest Statement

The authors have declared no competing interest.

### Author Declarations

All participants provided informed, written consent. A favourable ethical opinion for the main Genes & Health research study was granted by NRES Committee London - South East (reference 14/LO/1240) on 16 Sept 2014. Queen Mary University of London is the Sponsor, and Data Controller.

