## Supplementary table 1 for "The contribution of Apoliproprotein E genetic variation to dementia risk in British South Asians"

**Supplementary table 1: demographic characteristics.**

|  | Dementia | Controls |
| --- | --- | --- |
| **N** | 614 | 50,490 |
| **Age at recruitment (median, [IQR])** | 71.3 [15.9] | 39.0 [17.5] |
| **Gender (n, %)** |  |  |
| Female | 254 [41.4%] | 28,018 [55.5%] |
| Male | 260 [58.6%] | 22,472 [44.5%] |
| **APOE genotype** |  |  |
| ε2/ε2 | <10 [<1.7%] | 151 [0.3%] |
| ε2/ε3 | 42 [6.8%] | 3,794 [7.5%] |
| ε2/ε4 | <10 [<1.7%] | 475 [0.9%] |
| ε3/ε3 | 412 [67.1%] | 36,710 [72.7%] |
| ε3/ε4 | 131 [21.3%] | 8,776 [17.4%] |
| ε4/ε4 | 20 [3.3%] | 584 [1.2%] |
| **APOE allele frequency** |  |  |
| ε2 | 4.2% | 4.5% |
| ε3 | 81.2% | 85.2% |
| ε4 | 14.6% | 10.3% |
| **Inferred genetic ancestry** |  |  |
| Bangladeshi | 335 [54.6%] | 29,614 [58.7%] |
| Pakistani | 279 [45.4%] | 20,876 [41.3%] |
| **Age at diagnostic code (median, IQR)** | 73.3 [16.0] |  |
